# Fluoroquinolone-resistant *Escherichia coli* carriage in transrectal prostate biopsy patients without infectious complications

**DOI:** 10.1101/2022.08.15.22278533

**Authors:** Sofia Kalinen, Heini Kallio, Juha Knaapila, Teemu Kallonen, Eveliina Munukka, Tarja Lamminen, Pentti Huovinen, Peter J. Boström, Antti J. Hakanen, Marianne Gunell

## Abstract

Fluoroquinolones are a commonly used prophylaxis in transrectal ultrasound-guided prostate biopsy (TRUS-Bx), even though fluoroquinolone-resistant *Escherichia coli* has been associated with infectious complications after TRUS-Bx. The present study describes fluoroquinolone resistance mechanisms and antimicrobial susceptibility among intestinal *E. coli*, isolated from TRUS-Bx patients in a prospective study showing very few infectious prostate biopsy adverse events. This Multi-IMPROD sub-study included a total of 336 patients who received either ciprofloxacin, levofloxacin, or fosfomycin as prophylaxis before TRUS-Bx. *E. coli* could be cultured from 278 fecal swab samples, and 27 (9.7%) of these showed resistance to ciprofloxacin, and 14 (5.0%) were susceptible with increased exposure (I). Chromosomal and transferable fluoroquinolone resistance mechanisms were found among ciprofloxacin non-susceptible isolates, but both *qnr* genes and single *gyrA* mutations were found also among the ciprofloxacin-susceptible *E. coli* population. Low-level fluoroquinolone resistance is commonly associated with ESBL production in *Enterobacterales*. However, ESBL and *qnr* genes were not associated in our material, 14 isolates were ESBL producers and only 14.3% of them had the *qnr* gene, although 85.7% of the ESBL producers were ciprofloxacin non-susceptible. In the Multi-IMPROD substudy, only two mild urinary tract infections were reported, indicating that the antimicrobial susceptibility or resistance pattern of *E. coli* does not correlate with the onset of post-biopsy adverse events. We conclude that in our clinical settings, ciprofloxacin and levofloxacin prophylaxis is effective, and no severe post-biopsy infections were detected despite the intestinal colonization of genotypically and phenotypically fluoroquinolone-resistant *E. coli*.

## Introduction

Prostate cancer is one of the most common cancer types in men, especially in Western countries. A prostate cancer diagnosis is based on transrectal ultrasound-guided prostate biopsy (TRUS-Bx) which is an invasive procedure, and antimicrobial prophylaxis is a common practice to lower the risk for infectious adverse events. Fluoroquinolones are widely used prophylaxis in prostate biopsy procedures, due to their good penetration to prostate tissue (1). Fluoroquinolone resistance has emerged globally during the last decades, and both fluoroquinolone-resistant *Escherichia coli* and isolates with reduced susceptibility have increasingly reported to cause sepsis or other infectious adverse events after prostate biopsy procedure (2-6). In Finland, fluoroquinolones are very common per oral treatment for urinary tract infections in men. Fluoroquinolone resistance in both urine and blood *E. coli* isolates has increased during the last decade, being 15.1% and 10.8%, respectively in Finres data of 2020 (7). The percentage of ESBL-producing *E. coli* has also increased during the last decade being 6.5% in men’s urine isolates and 6.6% in blood isolates in 2020 (7). In *Enterobacterales*, low-level fluoroquinolone resistance has been associated with ESBL production (8-11), and in Finland, 2.3% of the urine *E. coli* isolates were both ESBL producers and fluoroquinolone-resistant in 2020 (7). Low-level fluoroquinolone resistance is mainly caused by plasmid-mediated quinolone resistance (PMQR) determinants like *qnr* genes whereas high-level resistance is caused by several chromosomal mutations in the quinolone resistance determining region (QRDR) of DNA gyrase (*gyrA/gyrB*) and topoisomerase IV genes (*parC*/*parE*) (12-14). In *E. coli*, resistance to fluoroquinolones is highly associated with mutations in *gyrA* (12). In the present study, we determined the correlation between prostate biopsy adverse events, fluoroquinolone resistance mechanisms and antimicrobial susceptibility among intestinal *E. coli* isolated from fecal swab samples from men undergoing TRUS-Bx procedure in Finland.

## Methods

### Study population

This study is a substudy of IMPROD (Improved Prostate Cancer Diagnosis – Combination of Magnetic Resonance Imaging (MRI) and Biomarkers, NCT02241122) multi-center study (15). A prostate cancer screen with TRUS-Bx was performed for patients included in this study and a rectal swab sample was taken during the biopsy procedure. Between March 2015 and May 2017, a total of 336 rectal swab samples were collected from four hospitals in Finland: Helsinki University Hospital, 58 samples; Tampere University Hospital, 59 samples; Satakunta Central Hospital, 87 samples; and Turku University Hospital, 132 samples. Levofloxacin was used as antimicrobial prophylaxis in Turku, and ciprofloxacin in all the other study sites. In Helsinki, patients who had traveled abroad within three months before the TRUS-Bx received fosfomycin instead of ciprofloxacin.

### Bacterial cultures

Swab samples were cultured on ChromAgar Orientation plate (Becton Dickinson, Heidelberg, Germany) and 5 µg-ciprofloxacin disk (OXOID, Thermo Scientific, Helsinki, Finland) was placed on top of the culture to select the patient’s most resistant *E. coli* strain. After the overnight incubation at 35 °C, two to three bacterial colonies with *E. coli* morphology (mauve to light purple colonies) were selected preferably near to the ciprofloxacin disk and pure cultures were made from these on CLED plates (Becton Dickinson, Heidelberg, Germany). Maldi-TOF (Bruker, Berlin, Germany) was used for species identification of the isolated strains. Only *E. coli* isolates were studied further.

### Antimicrobial susceptibility testing

Antimicrobial susceptibility testing was performed with the disk diffusion method, according to EUCAST guidelines (16). Following antimicrobial disks were used: ciprofloxacin 5 μg, levofloxacin 5 μg, cefotaxime 5 μg, ceftazidime 10 μg, cefoxitin 30 µg, meropenem 10 µg, ampicillin 10 µg, amoxicillin-clavulanic acid 10/20 µg, mecillinam 10 µg, nitrofurantoin 300 µg, trimethoprim 5 µg, and trimethoprim-sulfamethoxazole 25 µg (OXOID, Thermo Scientific, Helsinki, Finland). Antimicrobial susceptibility profiles of each *E. coli* isolates were determined according to EUCAST clinical breakpoint table version 11.0 2021 (17). *E. coli* ATCC 25922 was used as a control strain in antimicrobial susceptibility testing.

### PCR amplification and sequencing of chromosomal gyrA and parC gene mutations

Fluoroquinolone resistance mechanisms were studied in all *E. coli* isolates with a ciprofloxacin (CIP) disk inhibition zone ≤30mm. PCR amplification of *gyrA* and *parC* genes were performed with primers described in Table 1. The *gyrA/parC* PCR reaction (50 µL) consisted of 0.2 pmol/µL of each primer, 0.03 U/µL AmpliTaq Gold DNA polymerase, 5 µL AmpliTaq Gold buffer, 2 mM MgCl_2_ (Thermo Fisher Scientific Oy, Vantaa, Finland), and 0.2 mM dNTP mix (Life Technologies Europe, Espoo, Finland). The PCR program consisted of an initial denaturation at 94°C for 10 minutes, then 37 cycles of DNA denaturation at 94°C for 30 seconds, primer annealing at 55°C for 30 seconds, and extension at 72°C for 90 seconds. PCR-products were purified enzymatically with Exonuclease I- and FastAp Thermosensitive alkaline phosphatase -enzymes (Thermo Fisher Scientific Oy, Vantaa, Finland), and sequenced with BigDye v.3.1 sequencing using ABI3730xl DNA Analyzer at Institute for Molecular Medicine (FIMM, Helsinki, Finland). *Detection of plasmid-mediated quinolone resistance genes*. Transferable plasmid-mediated quinolone resistance (PMQR) *qnr* genes were screened for all the *E. coli* isolates with a ciprofloxacin disk inhibition zone ≤30mm. In addition, 34/90 randomly selected isolates with a ciprofloxacin inhibition zone >30mm were studied with previously reported primers and protocols (18).

**Table 1.**
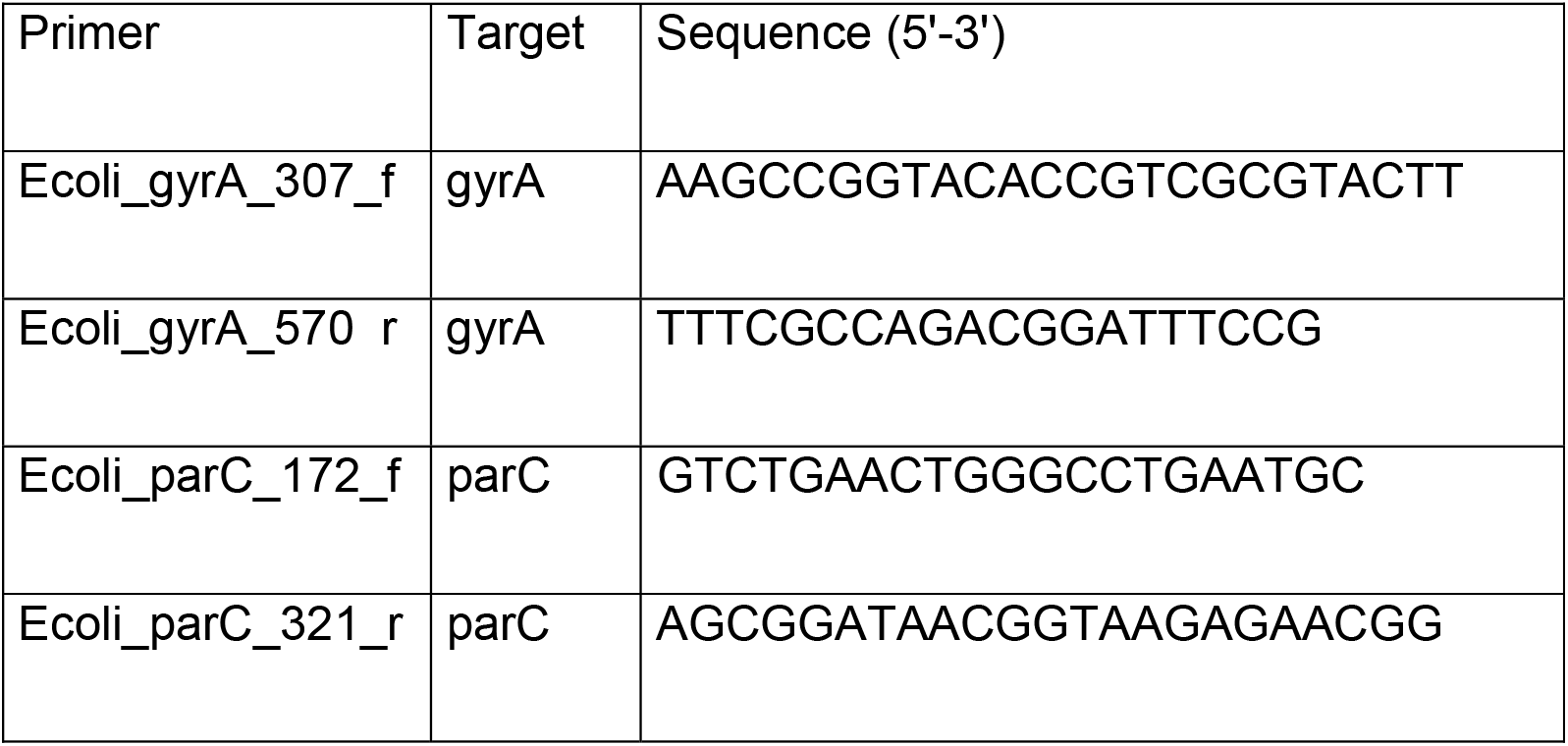
Primers used in *gyrA* and *parC* PCR and Sanger sequencing

### Detection of ESBL genes

Possible ESBL producers were screened according to EUCAST guidelines on the detection of Resistance mechanisms (19). ESBL + AmpC confirmation Kit (Rosco Diagnostics) was used to confirm ESBL production from *E. coli* isolates with reduced cefotaxime and ceftazidime susceptibility, inhibition zone <21 and <22mm, respectively. ESBL genes (*bla*_CTX-M,_ *bla*_SHV_ and *bla*_TEM_) were screened from these same isolates using previously described primers and PCR protocols (20).

## Results

### Antimicrobial susceptibility profiles

*E. coli* could be isolated from 278 patient samples (out of 336 fecal samples). The highest resistance rates were detected against ampicillin and amoxicillin-clavulanate (27.7% and 24.1%, respectively), also trimethoprim resistance was quite common (14.0%) (Figure 1). Totally 27 (9.7 %) *E. coli* strains were ciprofloxacin-resistant i.e. disk inhibition zone was <22mm and 14 (5.0 %) strains were ciprofloxacin-susceptible with increased exposure (I, CIP inhibition zone 22-24mm), according to the 2021 EUCAST breakpoints (17). Furthermore 22 (7.9%) *E. coli* strains were levofloxacin-resistant (R) (Figure 1). With correlation to given prophylaxis, 11 (9.6%) isolates from the levofloxacin group were resistant to both ciprofloxacin and levofloxacin, and six (5.3%) and four (3.5%) were ciprofloxacin- and levofloxacin-susceptible with increased exposure (I), respectively. Among the ciprofloxacin group, 16 (9.8%) and 11 (6.7%) isolates were ciprofloxacin and levofloxacin R, respectively, and eight (4.9%) and 11 (6.7%) isolates were ciprofloxacin and levofloxacin I, respectively.

**Figure 1.**
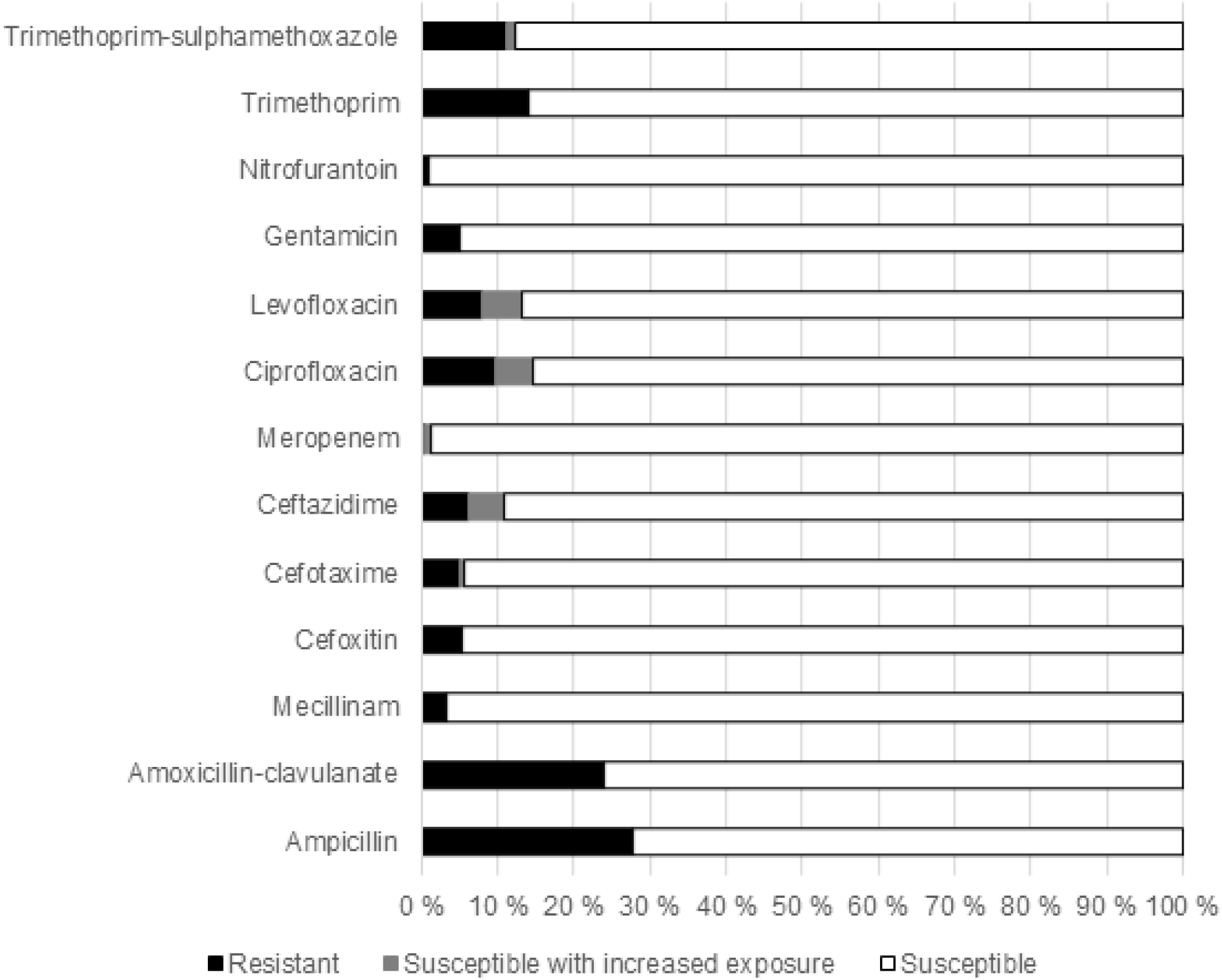
Antimicrobial susceptibility profiles of 278 *E. coli*, isolated from fecal swab samples taken from men undergoing TRUS-Bx

Resistance against cefotaxime and ceftazidime was detected in 14 (5.0%) and 13 (4.7%) *E. coli* isolates. Of the ciprofloxacin-resistant *E. coli* isolates, nine (3.2%) were cefotaxime-resistant, and eleven (4.0%) showed co-resistance to trimethoprim. These eleven strains were also resistant to ampicillin and sulfamethoxazole-trimethoprim. Four of these were also gentamicin-resistant and three of these were ESBL producing strains. Ten ciprofloxacin-resistant *E. coli* (3.6%) were also ESBL producers and five (1.8%) isolates were both trimethoprim-resistant and ESBL producers. Antimicrobial susceptibility results from all tested antimicrobials are presented in Figure 1.

### Fluoroquinolone resistance mechanisms and ESBL producers

Fluoroquinolone resistance mechanisms were found in all fluoroquinolone-non-susceptible isolates, i.e. CIP disk inhibition zone <25mm (17). Of the ciprofloxacin-resistant isolates, 23 had point mutations in the QRDR of both *qyrA* and *parC*, and four had point mutations only in *gyrA*. The most common *gyrA* mutation was S83L + D87N –double mutation found in 20 isolates, nine of these had also S80I + E84G –double mutation, and ten had S80I –single mutation in *parC* (Figure 2). Of the 14 *E. coli* isolates being ciprofloxacin I (susceptible with increased dose, according to EUCAST (17), disk inhibition zone 22-24mm), ten had S83L –single mutation and one had D87Y –single mutation in the QRDR of *gyrA*, and three isolates had only *qnr* gene (*qnrA/B*) as their only fluoroquinolone-resistance mechanism detected.

**Figure 2.**
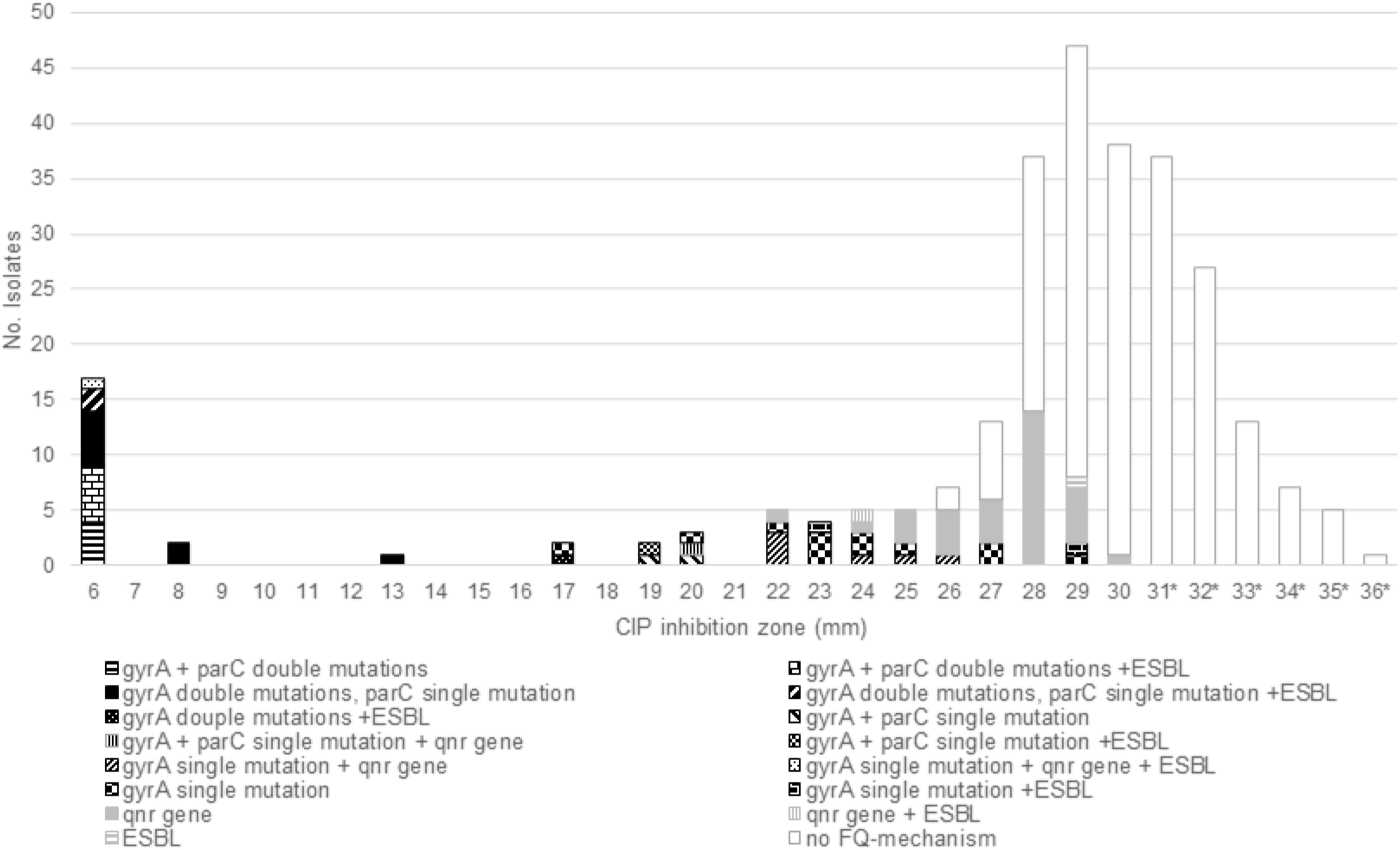
Fluoroquinolone (FQ) resistance mechanisms and ESBL genes detected among 276 *E. coli* isolates with ciprofloxacin (CIP) disk inhibition zone 6-30 mm. A dashed black line with R represents EUCAST and CLSI breakpoint (16, 23) for CIP-resistant isolates. A dashed line with black S represents the EUCAST breakpoint and a grey dashed line with S represents the CLSI breakpoint for CIP susceptible isolates. R= resistant; S= susceptible *Randomly selected 34 isolates with disk inhibition zone 31-36mm were tested for *qnr* genes

Fluoroquinolone resistance mechanisms were also found among ciprofloxacin-susceptible isolates. A total of seven isolates with ciprofloxacin inhibition zone between 25-29mm had a single *gyrA* mutation, and 31 isolates had *qnr* gene. The distribution of fluoroquinolone resistance mechanisms among the tested *E. coli* population is presented in Figure 2.

According to EUCAST Detection of resistance mechanisms guideline (19), 15 *E. coli* isolates (5.4%) were ESBL producers, nine of these were positive for *bla*_CTX-M_ gene, four for *bla*_CTX-M_+*bla*_TEM_, and one strain for *bla*_SHV_ +*bla*_TEM_. One screening positive isolate did not have any of these three ESBL genes analyzed. Of the ESBL *E. coli* strains, ten were ciprofloxacin-resistant and had chromosomal mutations, two were ciprofloxacin susceptible with increased dose, one of them had *qnrS* gene and the other had a single *gyrA* mutation. Two of the ESBL strains were ciprofloxacin-susceptible, but one of these had a single *gyrA* mutation. The distribution of ESBL genes and fluoroquinolone resistance mechanisms is presented in Figure 2.

### Fluoroquinolone susceptibility and resistance mechanisms in correlation to post-biopsy adverse events

In the Multi-IMPROD study, only 12 minor post-biopsy adverse events were reported.^21^ The list of complications, prophylaxis, and microbiological findings, including ciprofloxacin susceptibility and possible resistance mechanisms of these 12 patients’ are presented in Table 2.

**Table 2.** Patients with post-biopsy adverse events, used prophylaxis, ciprofloxacin susceptibility testing results and resistance mechanisms detected.

| Sample ID | Prophylaxis | Adverse event | E. coli | CIP disk inhibition zone (mm) | Susceptibility interpretation | FQ-resistance mechanism |
| --- | --- | --- | --- | --- | --- | --- |
| TA-009 | Ciprofloxacin | mild UTI | yes | 24 | I | S83L |
| TU-062 | Levofloxacin | hematuria | yes | 27 | S | D87N |
| HE-056 | Ciprofloxacin | hematuria | yes | 28 | S | *qnrA* |
| PO-071 | Ciprofloxacin | urine retention | yes | 28 | S | - |
| PO-055 | Ciprofloxacin | mild UTI | yes | 29 | S | - |
| TU-010 | Levofloxacin | hematuria | yes | 30 | S | - |
| TU-054 | Levofloxacin | hematuria | yes | 30 | S | - |
| HE-053 | Ciprofloxacin | hematuria | yes | 31 | S | - |
| PO-075 | Ciprofloxacin | urinary retention | yes | 31 | S | - |
| TU-146 | Levofloxacin | rectal bleeding | yes | 31 | S | - |
| TU-026 | Levofloxacin | hematuria | yes | 32 | S | - |
| HE-047 | Ciprofloxacin | hematuria/<br>urinary<br>retention | no | nd | nd | nd |

## Discussion

In the present study, the antimicrobial susceptibility and fluoroquinolone resistance mechanisms of intestinal *E. coli*, isolated from faecal swab samples from men undergoing prostate biopsy procedures were analysed. Post-biopsy infections are a common side effect of prostate biopsy procedures, and especially fluoroquinolone-resistant and multidrug-resistant *E. coli* has been reported of being the main factor for infectious adverse events (2-5, 22). Our results showed that 14.7% of the tested *E. coli* strains were ciprofloxacin non-susceptible, and 85.3% were susceptible according to EUCAST breakpoints (17). If CLSI breakpoints would have been used the number of ciprofloxacin-resistant strains would have been the same (CIP ≤21mm), whereas the ciprofloxacin I isolates would have increased from 14 to 19 isolates (CIP 22-25mm vs. CIP 22-24mm) (23). Resistance to levofloxacin was more common in the study population who got levofloxacin prophylaxis than in ciprofloxacin prophylaxis group (9.6% vs. 6.7%, respectively), whereas ciprofloxacin resistance was practically equal in both groups (9.6% vs. 9.8%, respectively). Of the ciprofloxacin-resistant *E. coli*, 4.0% showed co-resistance to trimethoprim, 3.6% of were ciprofloxacin-resistant and ESBL producers, and 1.8% were ciprofloxacin- and trimethoprim-resistant and ESBL producers. Compared to overall susceptibility levels of *E. coli* isolates in Finland in 2020, fluoroquinolone resistance among the study population was higher compared to invasive isolates (10.8%), but equal to resistance levels in urine isolates (15.1%) (7). ESBL percentage among the intestinal *E. coli* isolates was somewhat higher, 5.4%, compared to the earlier reported ESBL carriage rate (4.7%) in Finland (24), but lower compared to ESBL-positivity rate in urine *E. coli* – isolates (6.5%) (7). In addition, resistance to trimethoprim was lower, whereas multidrug resistance was more common compared to that reported in Finres 2020 (7).

To become a high-level fluoroquinolone-resistant, bacterial strain needs to have mutations in both QRDR of *gyrA* and *parC*, however mutation even in one of these gene targets, S83L in *gyrA*, S80I in *parC*, and D87N in *gyrA* can cause a clinically relevant fluoroquinolone resistance in *E. coli* (25). In our study, double *gyrA*+*parC* mutations were detected only in high-level resistant isolates whereas single *gyrA* mutations were detected even phenotypically ciprofloxacin-susceptible isolates. EUCAST has started using the term ATU (Area of Technical Uncertainty) to warn laboratories that, there can be interpretative difficulties with susceptibility test results in this area, and susceptibility interpretation should be carefully evaluated (26). According to EUCAST breakpoints for ciprofloxacin (17), the ATU area is situated between the resistant and susceptible populations (22-24mm), i.e. I area, and includes the isolates which can be considered susceptible with increased exposure. As we show in Figure 2, isolates within the area of technical uncertainty, have chromosomal gyrase mutations, indicating that it is justified to consider these isolates rather resistant than susceptible. PMQR determinants like *qnr* genes are linked to low-level fluoroquinolone resistance but these genes also enhance the selection of high-level resistance, furthermore, the *qnr* genes are also easily missed until further mechanisms are acquired and detected (11-12,14, 27-28). We show that the *qnr* genes were detected among ciprofloxacin-resistant, ciprofloxacin I, and ciprofloxacin-susceptible strains, indicating that *qnr* genes do not necessarily have a clinical relevance without additional resistance mechanisms (25). PMQR genes are associated with the same plasmids as ESBL genes (9-11, 27) and there are reports on post-biopsy infections linked to ESBL *E. coli* isolates with co-resistance to fluoroquinolones (29). In our study, 85.7% of the ESBL strains had chromosomal mutations in QRDR and only one ESBL strain had a PMQR gene as the only fluoroquinolone resistance mechanism, indicating that ESBL-producing strains and strains with *qnr* genes were not associated in our material. In addition, among the tested ciprofloxacin-susceptible *E. coli* isolates with disk inhibition zone 31-36mm, no fluoroquinolone resistance determinants were detected, as expected. Despite the ciprofloxacin and multidrug resistance detected in our study population, no severe post-biopsy infections were reported in the Multi-IMPROD study (21). It is known, that the use of fluoroquinolones increases the risk for intestinal colonization of fluoroquinolone-resistant *E. coli*, and further post-biopsy infectious complications (6,30). In the present study, there were both phenotypically ciprofloxacin-resistant *E. coli* and isolates with susceptible phenotype but having fluoroquinolone resistance mechanisms, and despite this, only two mild UTI cases were reported. One of them had ciprofloxacin-susceptible *E. coli* in a swab sample, and the other had ciprofloxacin I *E. coli* with a single *gyrA* mutation, indicating that fluoroquinolone resistance alone does not explain the possible adverse events after TRUS-Bx. There are reports on certain *E. coli* clones, such as ST131 that have spread widely and have caused severe infections (3), and thus further studies are needed to evaluate the role of virulence factors and other resistance mechanisms such as efflux pumps, in post-biopsy infections.

## Conclusion

We show in this study that fluoroquinolone resistance mechanisms, both the *qnr* genes and *gyrA* mutations, were also found among the ciprofloxacin-susceptible *E. coli* population and that ESBL and transferable *qnr* genes were not associated in our material. We conclude that the onset of post-biopsy adverse events did not correlate with antimicrobial susceptibility, since no severe post-biopsy infections were detected despite the intestinal colonization of genotypically and phenotypically fluoroquinolone-resistant *E. coli*.

## Data Availability

All data produced in the present study are available upon reasonable request to the authors

## Acknowledgments

We thank Minna Lamppu for her skillful technical assistance with cultured *E. coli* isolates

## Transparency declarations

None to declare

